# Antibiotic use for typhoid in India: Estimation from private sector prescription data during 2013-15

**DOI:** 10.1101/2021.12.10.21267591

**Authors:** Shaffi Fazaludeen Koya, Habib Hasan Farooqui, Aashna Mehta, Sakthivel Selvaraj, Sandro Galea

## Abstract

**Background:** India’s typhoid burden estimates are based on a limited number of population-based studies and data from a grossly incomplete disease surveillance system. In this study, we estimated the total and sex- and age-specific antibiotic prescription rates for typhoid.

**Methods:** We used systematic antibiotic prescription by private sector primary care physicians in India. We categorized antibiotics using the WHO classification system and calculated the prescription for various classes of antibiotics.

**Results:** We analyzed 671 million prescriptions for the three-year period (2013-2015), of which an average of 8.98 million antibiotic prescriptions per year was for typhoid, accounting for 714 prescriptions per 100,000 population. Combination antibiotics are the preferred choice of prescribers in the adult age group, while cephalosporins are the preferred choice in children and young age. The prescription rate decreased from 792/100,000 in 2013 to 635 in 2015.

**Conclusion:** We report a higher rate of antibiotic prescription for typhoid using prescription data, indicating a higher disease burden than previously estimated. Quinolones are still widely used in monotherapy, and children less than 10 years account for more than a million cases annually, which calls for a routine vaccination program.

**What we already know:**

- Typhoid is a major cause of morbidity in India especially among young adults and children.
- The reported incidences are based on data from limited number of population-based studies and the disease surveillance program which is largely limited to public healthcare system in India.
- The emergence of antibiotic resistance among typhoid is a growing concern.

**What this article adds:**

- Age-specific typhoid antibiotic prescription estimate for India, using a large volume of geographically representative medical audit data.
- We report a high rate of antibiotic prescription (714/100,000 population) for typhoid indicating a higher disease burden than previously estimated.
- Fluroquinolones are still widely used as monotherapy for the treatment of typhoid in India.

## Introduction

Enteric Fever, a systemic infection caused by Salmonella enterica serotypes *S. typhi* and *S. para typhi*, remains an important public health problem.^1^ World Health Organization recommends vaccination as a control strategy in the short to intermediate terms.^2,3,4^ Globally, there was a 44% decline in typhoid and para-typhoid fever between 1990 and 2017.^5^ With improved water hygiene and sanitation, and an increase in vaccine uptake, water-borne diseases have declined in general.

Although India’s disease surveillance system includes typhoid, there are several challenges to assess the burden of the disease.^6^ This includes poor or incomplete reporting by the private sector that caters to the majority of outpatient care in the country.^7^ The top ten causes of death in India for the year 2017 include communicable diseases, namely, diarrheal diseases, lower respiratory infections, and tuberculosis.^8^ India remains one of the high burden countries of typhoid. The global burden of diseases (GBD) 2017 typhoid and paratyphoid collaborators estimated the typhoid/para typhoid incidence as 586.3 per 100,000 population (95% UI: 515.7, 661.8)^5^ Similarly, a systematic review in 2016 estimated annual incidence of typhoid as 377/100,000 (95% CI: 178–801) and that of para-typhoid as 105/100,000 persons (95% CI: 74–148). ^9^

These reported incidences are based on data from a limited number of population-based studies and the disease surveillance system which is largely limited to the public healthcare system in India. These estimates are prone to the risk of either over estimation or under estimation due to non-uniformity in the definition and diagnostic methods adopted to detect typhoid disease and the limited sample size in the population-based studies. At the same time, the relatively easy availability of low-cost antibiotics without prescription leads to lower probability of diagnosis and reporting through formal healthcare, low rates of confirmatory diagnostic testing for typhoid, and low sensitivity of blood culture tests remain as challenges for effective typhoid surveillance in India.^10^ The world’s first Typhoid conjugate vaccine (Typbar TCV®) produced in India was introduced at the community level in 2018 in Mumbai on a pilot basis after WHO prequalification.^11^ It is important to estimate the disease burden to measure the impact of already ongoing health interventions like Swachh Bharat (clean India) mission^12^ and Jal Jeevan (drinking water) mission^13^ but also to decide on the inclusion of the typhoid vaccine in the immunization schedule.

The emergence of antibiotic resistance among typhoid is also a growing concern.^14,15^ Studies show that resistance to quinolones has increased in recent years, ampicillin and trimethoprim–sulfamethoxazole resistance have decreased, and resistance to third-generation cephalosporins remains low.^16^ Data on prescription and sales of antibiotics can be insightful to understand the burden of disease and variations across age groups, sex, and regions besides prescription patterns.^17,18,19^ We aimed to use the antibiotic prescription data from a large nationally representative survey of private sector general practitioners to understand the magnitude and age and sex-specific rates of antibiotic prescriptions for typhoid in India.

## Methods

We used data on systemic antibiotics (J01) prescription by private sector primary care physicians in India collected by IQVIA (formerly IMS Health) for the years 2013, 2014, and 2015.^20^ IQVIA collects data and provides information on medical practice, especially on the use of medicines in over 100 countries around the world. The monthly prescription audit data in India pertains to prescriptions by a panel of 4600 clinicians who practice modern medicine selected through a multistage stratified random sampling accounting for the region specialty type, and patient turnover. The sample includes general practitioners, specialist physicians, dentists, and AYUSH (non-MBBS doctors) from 23 metropolitan areas (population more than 1 million), 128 class 1 towns (population 100,000-1 million) and 1A towns (population less than 100,000). The data is then extrapolated to reflect the private sector prescription pattern.

This database provides information on patient characteristics such as age, gender, diagnosis, and medicines prescribed, besides the location details (zone-east, west, north, south) and urban locality (Metropolitan cities or class 1/1A towns). IQVIA organizes medicines according to the anatomical therapeutic classification (ATC) of the European Pharmaceutical Market Research Association (EphMRA), but the authors used the ATC index provided by the World Health Organization WHO collaborating center to convert them to the WHO ATC classification.^21^ [Table S1, Supplement]. We extracted the information on the diagnosis reported on prescriptions and used the ICD codes A01.0 and A01.10 to identify typhoid and para typhoid cases, respectively. We used the aggregated, processed, and extrapolated data to estimate the total antibiotic prescriptions for typhoid to understand the antibiotic prescription practices for typhoid in the country. We further used the India population data and the age structure of Indian population from the population pyramid to calculate sex and age-specific rates of antibiotic prescriptions for typhoid.^22^ For doing the age-specific analysis, we used only the prescriptions with data for age. In addition, we also tried to compare the prescription patterns with the available information on antibiotic resistance for typhoid for a selected classes of antibiotics for specific years.

In the private sector, antibiotics are usually prescribed for the entire duration of the course of treatment for a particular disease. In that sense, each prescription of an antibiotic corresponds to a diagnosed case of typhoid and therefore, it is a good proxy for measuring the prevalence of typhoid. However, the data does not capture the public sector prescriptions and therefore our analysis only reflects outpatient typhoid diagnosis and antibiotic prescription patterns in the private sector in the country.

## Ethical approval

Individual-level data was not collected and there was no personal identifier in the dataset that we analyzed. Therefore, we did not require ethical approval for our study.

## Results

We analyzed 671 million prescriptions for the three-year period (2013-2015), of which 26.96 million antibiotic prescriptions were made for enteric fever (typhoid and paratyphoid cases), averaging 8.98 million per year in in India. The prescriptions of enteric fever (typhoid and paratyphoid cases) decreased by 9.5% between 2013 and 2014 (from 9.9 million in 2013 to 9.1 million in 2014) and further by 11.3% to 7.9 million in 2015. The data was scanty for para-typhoid fever (only 1163 total cases in 2013, 315 in 2014, and 124 in 2015), and therefore the data largely represents typhoid fever in the country. Overall, typhoid cases were more among males. North and west regions of the country had the highest reported cases. Majority of cases were reported from metropolitan cities. Adolescents and young adults have the highest burden in absolute numbers, with close to 20% patients belonging to 10-19 age group. Another 13 to 14% patients are in the 5-9 and 20-29 age groups. [Table S2, Supplement]

The average annual countrywide antibiotic prescription rate for typhoid was 714/100,000 population during the period 2013-2015. The prescription rate varied across age groups and gender. Males had a higher average rate (844/100,000) compared to females (627/ 100,000) over the three-year period. Over the three-year period (2013-2015), the age groups 0-4 years and 10-19 years showed a similar average rate (479/100,000). However, 10-19 years age group represented 18.6% of the total burden in the country in absolute numbers. On average, more than 35% of the cases were below 20 years of age. [Figure 1], The rate decreased from 792/100,000 in 2013 to 716/100,000 in 2014 and further to 635/ 100,000 population in 2015. The rate among males decreased by 22% (947/100,000 in 2013 to 738/100,000 in 2015) and among females by 17% (683/100,000 in 2013 to 570/100,000 in 2015) over the same period. [Figure 2]. The number of prescriptions sharply increases in the age group 20-29 years (806/100,000). With more than a quarter (26.4%) of the total cases in the country, the 20–29-year age group also had the highest age-specific rate. The prescription rate decreases sharply after the age of 30. [Figure 3], There were clear differences in the number and rate of prescriptions between the sexes in all age groups, with males sharing a higher burden. The difference was maximum in the age group 0-4 years (28% higher for boys) while the age group 20-29 had the least difference (8%) [Table S3, Supplement] Combination antibiotics (J01R) are the preferred choice in the adult age group, while cephalosporins (J01D) are the preferred choice of prescribers in children and young age (up to 20 years). We didn’t observe any major changes in the prescription share for antibiotic classes over the three-year period. [Figure 4]

**Figure 1:**
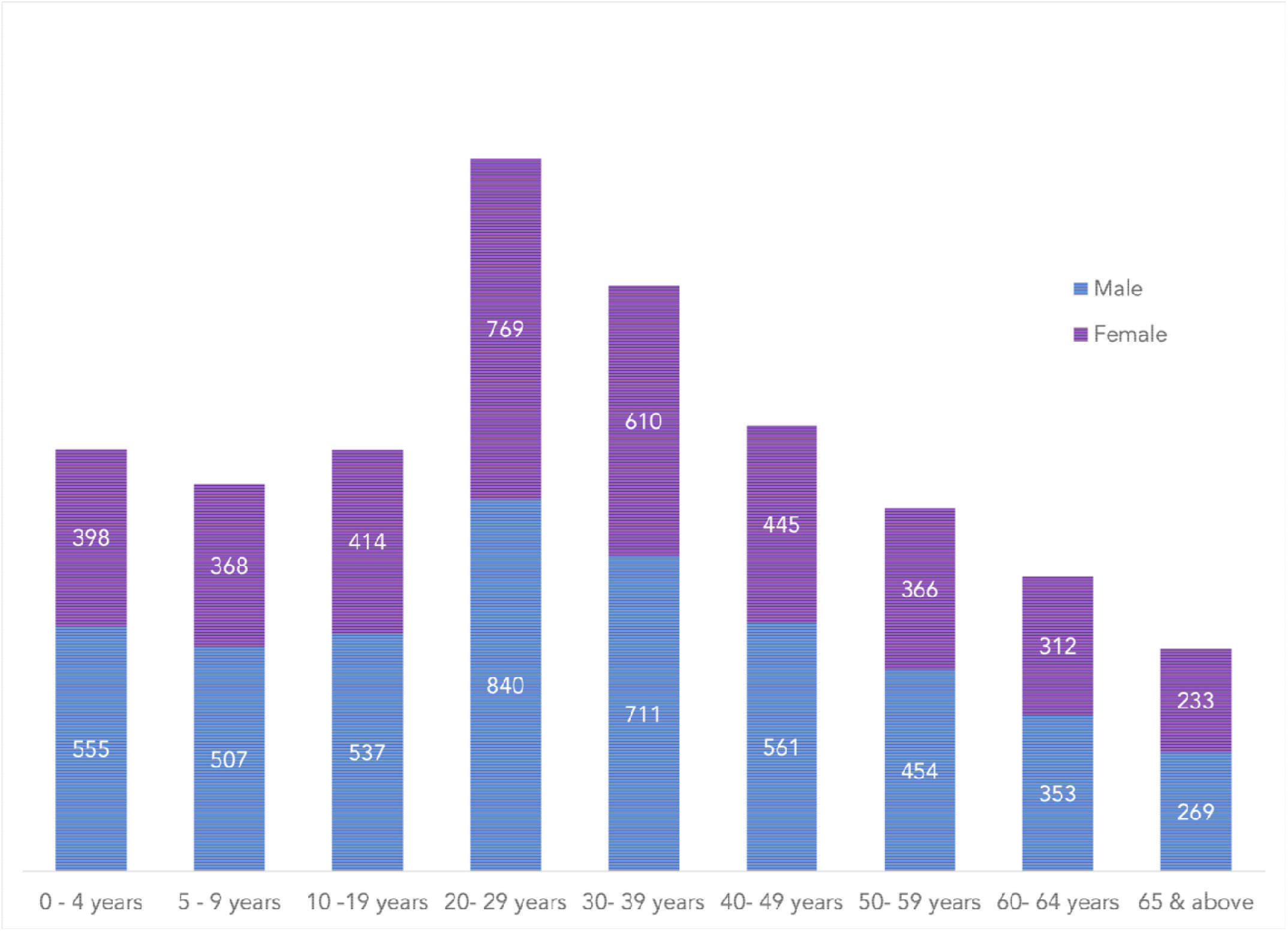
Three-year average antibiotic prescriptions (2013-15) for typhoid, across various age groups, per 100,000 population

**Figure 2:**
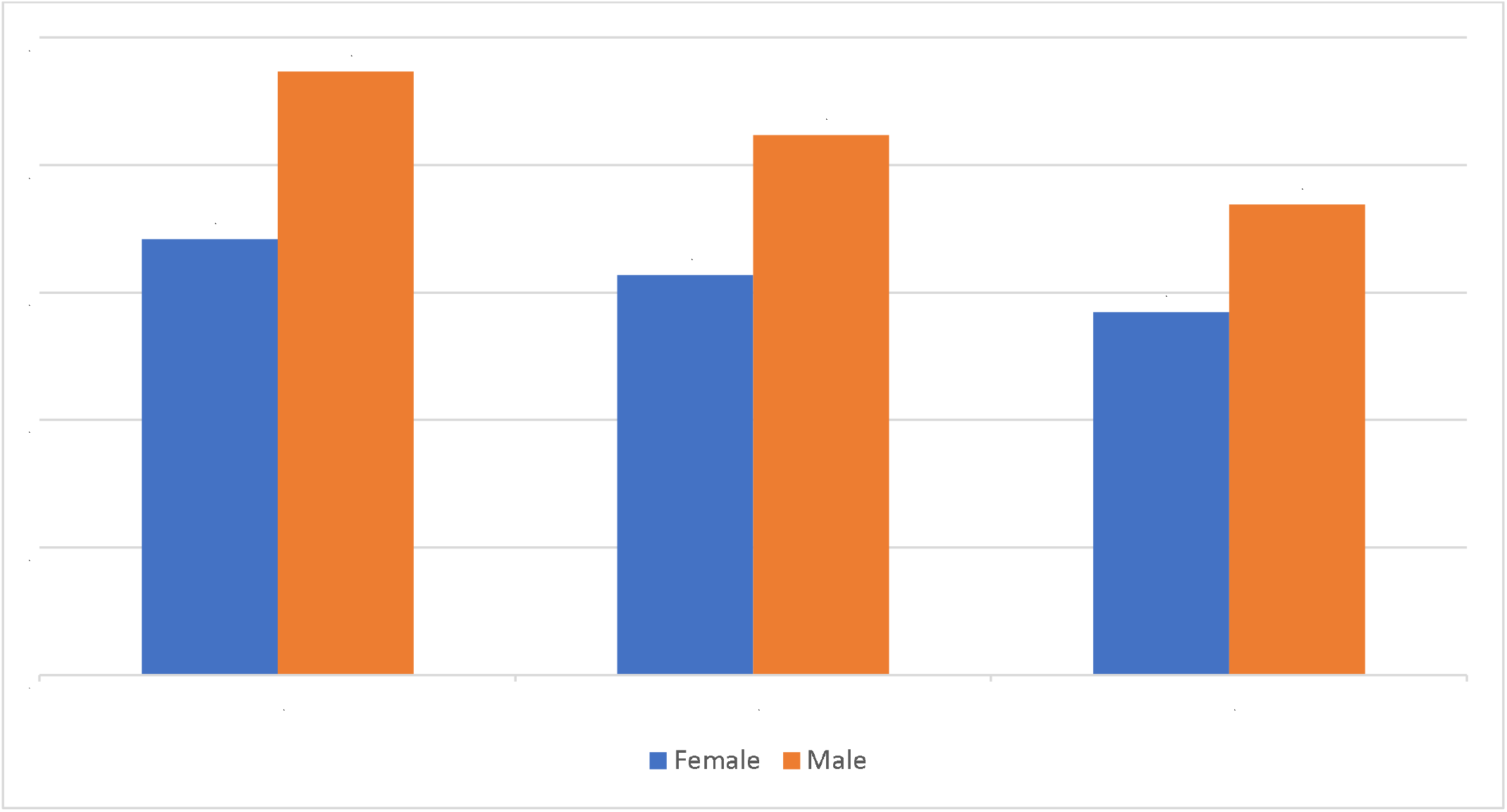
Annual age-adjusted, sex specific rates (per 100,000 population) for typhoid antibiotic prescriptions, 2013-2015

**Figure 3:**
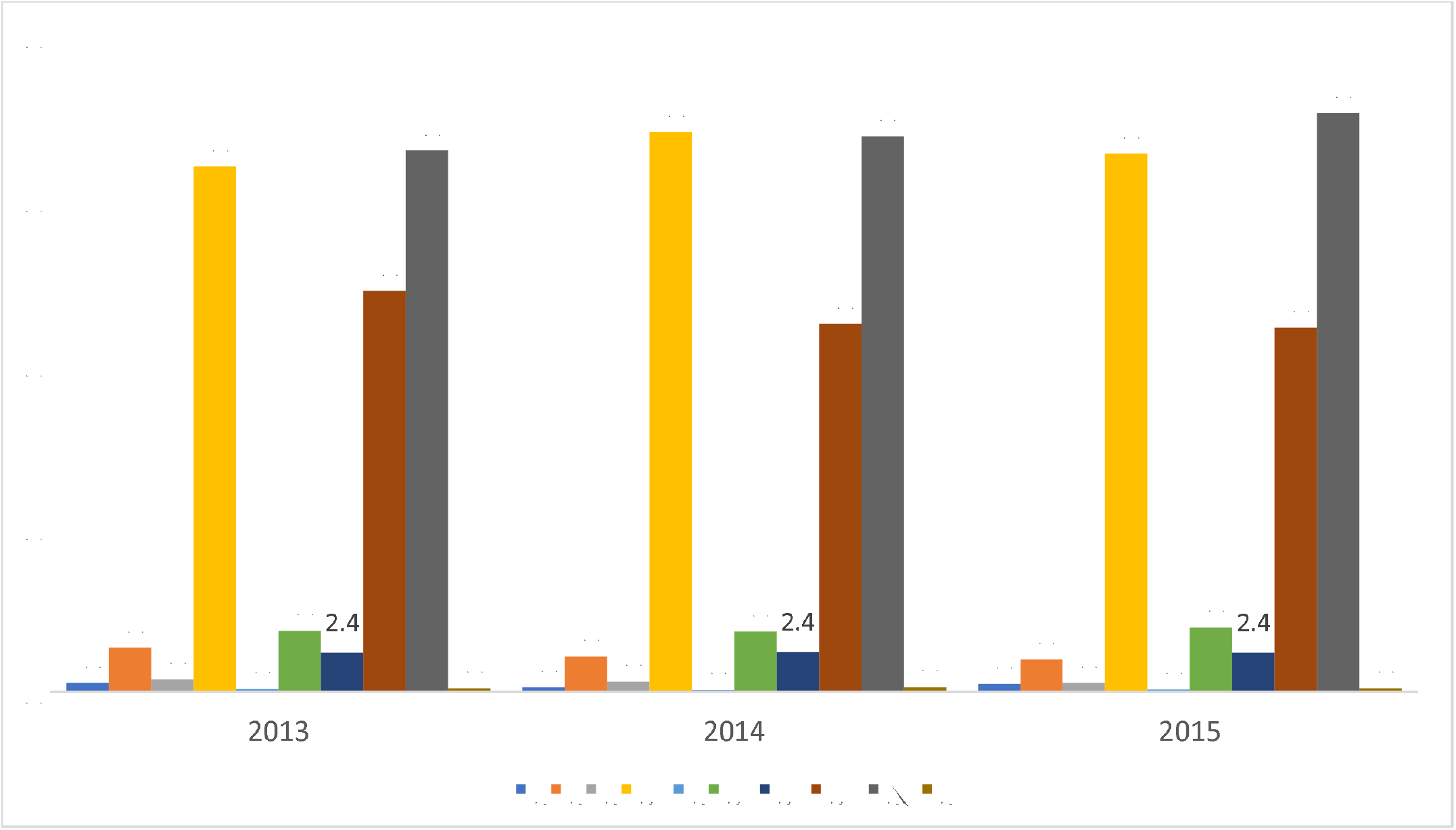
Share (in percentage) of various antibiotic classes for typhoid across years 2013 to 2015

On average, there were 108 different formulations of antibiotics prescribed for typhoid, ranging from 80 to 96 across geographical zones. Formulations varied across age groups, ranging from 63 in patients aged 65 years and above to 84 in 20-29 age group. Young adults were treated with a wide range of formulations. Ten different antibiotics accounted for three-quarter of all prescriptions (72.4%). Cefixime-Ofloxacin combination was the preferred drug of choice for typhoid across metro and class 1 cities and across regions except south India, where cefixime was the most prescribed antibiotic. (Figures S4-S7, Supplement). Ciprofloxacin is still widely used in west and south regions and in class 1/1A towns, whereas it was not among the top five preferred antibiotics in metro as well as north and east regions.

Combinations of antibiotics (mostly a combination of Cephalosporin and Fluroquinolone) and Cephalosporins are the most used antibiotic classes, both in metro cities and class 1/1A towns. Antibiotics in the WHO antibiotic class J01 R (“combination of two or more antibiotics”) were used to treat 33.9% typhoid patients, followed by J01 D (Cephalosporins) which were used in 32.9% of patients [Table S8, Supplement]. The age group wise preference of antibiotic class is given in Supplement [Figure S8].

## Discussion

To our knowledge, this is the first age-specific typhoid antibiotic prescription estimate for India, using a large volume of geographically representative medical audit data. This study reports a typhoid related average antibiotic prescription rate of 714/100,000 population during the three years (2013-2015), that signals higher estimate of typhoid burden in the country compared to some previous reports including a systematic review in 2016 which estimated an incidence of 377/100,000 and the GBD 2017 estimate of 586.3/100,000 population. ^5,9,23^ However, considering that our numerator includes only the population being catered by private practitioners and the denominator includes the whole population, this may still be an underestimate. Our study used data from private providers which caters for 70% of outpatient care in India ^19,24^. Outpatient record represent the majority of the typhoid related prescription as only six out of every 1000 typhoid cases require hospitalization^25,26,27^. Furthermore, long-term studies like the ongoing multisite study among pediatric cohorts^28^ will be helpful in generating more real world estimates on the burden, cost, and sequalae of the infection.

Resistance to typhoid antibiotics is a global public health issue.^15,29,30^ Antibiotic resistance in typhoid is a well-acknowledged problem in India as well.^31,32,33^ Available data shows that resistance to quinolones, the most commonly used class of antibiotics for typhoid, has been consistently increasing in India, from 11% in 2008 to 68% in 2015 whereas resistance to another most commonly used class, cephalosporins remained low.^34^ Resistance to the other classes of antibiotics ranges from 8% for penicillin to 12% for aminoglycosides, and to 23% for trimethoprim-sulfamethoxazole. Interestingly, a recent systematic review showed that the typhoid antibiotic resistance in India has moved from a multi-drug resistance pattern to one primarily led by quinolone resistance.^31^

Our study showed similarities and differences in antibiotic prescription preferences among practitioners across the four regions of the country. Our analysis shows that a combination of cephalosporins and quinolones is the preferred antibiotic of choice by providers in India. However, a significant proportion of cases in India are still treated with quinolones alone (23%), and the top five antibiotics used in the south and west regions of the country include ciprofloxacin and ofloxacin. We found that ofloxacin is the third most common antibiotic used. Ciprofloxacin is widely used as monotherapy, at least in the west and south regions, even though the drug was known to have resistance since two decades.^35^ WHO recommends ciprofloxacin as well as ofloxacin only for fully sensitive typhoid cases and in the absence of culture sensitivity tests for most of the typhoid cases diagnosed, the use of these drugs as monotherapy needs attention, especially in high endemic regions of the country.

In India, the highest proportion of hospitalization is still due to infections including typhoid.^36^ The cost of treating an episode of typhoid in outpatient care ranges from $2.0-$2.6 (mean, $2.3, 2010 US$), and from $96 to $132(mean $113, 2010 US$) for hospitalized care.^37^

With the availability of a new prequalified conjugate typhoid vaccine, a discussion around the introduction of a conjugate typhoid vaccine is required. Furthermore, recent literature suggests the vaccine to be cost effective as a preventive strategy for typhoid.^38^ Trials including those conducted in India have shown that the currently available conjugate vaccine is safe and highly immunogenic.^39,40^ Modelling based studies and clinical trials have also highlighted that the introduction of pathogen-specific vaccines reduces demand for antibiotics by reducing the force of transmission and incidence of diseases which consequently has an impact on the reduction of antibiotic resistance also.^41^

The age-specific rates in our study corroborates with some recent studies from Vietnam, Bangladesh, and Pakistan, besides Kolkata in India, all of which showed varying patterns of age-specific incidence of typhoid.^26, 41–45^ Contrary to earlier understanding^9^, our analysis shows that higher proportion of young adults than children are treated every a year for typhoid in India. This may be because of the atypical nature of the clinical presentation of typhoid in children, especially among less than 5 years, that might lead to reduced laboratory testing for typhoid among young age group and a subsequent smaller number of cases being diagnosed. Even then, our analysis suggests that around 2 million prescriptions in the year 2013, 1.8 million in 2014, and 1.1 million prescriptions in 2015 were issued for children less than ten years of age. This calls for well-designed prospective studies^27^ and community-level surveillance systems across various regions to understand the true age specific incidence in the Indian context.^28^

## Limitations

The study has a few limitations. The study used prescription data from a representative sample of private sector providers. The data does not have information on the laboratory confirmation of typhoid and therefore some degree of misclassification can be expected. However, this is reflective of the real-world setting where laboratory confirmation is not the norm. The prescription data pertains to private sector providers in small towns and urban areas. However, there is not much reason to believe that the prescription will be different in rural areas, although the prescription patterns may be different in the public sector. Finally, we excluded 20% prescriptions from age-specific analysis as they didn’t have data on age groups, which might have underestimated the age-specific rates in some age groups and overestimated in some others.

## Conclusion

Using a large volume of private sector data, we found that typhoid antibiotic prescription in India has decreased by 2 million between 2013 and 2015. Still, the country has a large burden of typhoid with 7.9 million prescriptions in 2015, corresponding to around 635 typhoid cases/million population. There is variation in antibiotic usage across ages and regions. Quinolones are still widely used in monotherapy, despite evidence of high resistance. Young patients account for close to one-third of the cases and children less than 10 years account for more than a million cases annually. Routine vaccination programs alongside improvement in water, hygiene and sanitation facilities can help to decrease the typhoid cases drastically.

## Data Availability

The data underlying the results presented in the study are available from IQVIA.(www.iqvia.com)

